# SARS-CoV-2, the human placenta, and adverse perinatal outcomes

**DOI:** 10.1101/2024.09.20.24313999

**Authors:** Rebecca L. Linn, Markolline Forkpa, Rita Leite, Jessenia C. Guerrero, Maria C. Reyes, Lauren E. Schwartz, Rebecca A. Simmons, Samuel Parry, Thea N. Golden

## Abstract

The relationship among timing and severity of COVID-19 during pregnancy, placental pathology, and adverse pregnancy outcomes is not well understood. We conducted a prospective cohort study of 497 pregnant patients with COVID-19 whose placentas underwent systematic pathologic examination. The main exposure was timing of COVID-19 during pregnancy (first or second versus third trimester). The primary outcome was composite placental pathology that included high grade maternal vascular malperfusion or greater than 25 percent perivillous fibrin deposition. There were 63 patients who had the composite placental pathology outcome. In adjusted analyses that controlled for maternal age, parity, active infection at delivery, interval from time of diagnosis to delivery and COVID-19 variant, timing of COVID-19 during pregnancy was not associated with risk of the composite placental pathology outcome. Among secondary COVID-19 related exposures we investigated, severity of disease and treatment for COVID-19 were associated with risk of the composite placental pathology outcome. In addition, patients with COVID-19 in the first nine months of the pandemic had the highest rate of the composite placental pathology outcome. In this large cohort, placental vascular pathology was common among COVID-19 cases but was unrelated to timing of COVID-19 during pregnancy or adverse pregnancy outcomes. These findings suggest that uncomplicated COVID-19 during pregnancy does not require intensive fetal surveillance or detailed pathologic examination of the placenta after delivery.

## Introduction

Pregnancy appears to be a risk factor for severe disease with 2019 novel coronavirus disease (COVID-19). Immediately following the start of the pandemic case studies were published reporting the increased incidence of maternal morbidity and mortality and adverse pregnancy outcomes. We now have a better understanding of the increased risk of adverse outcomes following COVID-19 during pregnancy.^1–4^ Smith et al. recently published a meta-analysis that included over 13,000 pregnancies included in 12 studies across 12 countries.^3^ They found a significantly greater risk of mortality, admission to intensive care unit, and other morbidities among pregnant patients with COVID-19. They also found an increase in preterm birth and low birth weight if the neonate was born to a patient with severe acute respiratory syndrome coronavirus (SARS-CoV-2) infection during pregnancy. Many other reports associate COVID-19 with adverse pregnancy outcomes, including fetal growth restriction, hypertensive disorders of pregnancy, and fetal loss. ^3,5–7^ Most of these adverse pregnancy outcomes are attributed to placental dysfunction,^8–12^ but the effects of COVID-19 on placental pathology and placental function have not been fully elucidated.^13–17^

Placental pathology frequently is reported in pregnancies complicated by maternal SARS-CoV-2 infection. Vasculopathies and inflammation have been reported commonly in case series and small cohort studies.^13,14,17–21^ Corbetta-Rastelli et al. recently conducted a relatively large cohort study that included 138 placentas with infections during pregnancy.^22^ Similar to many smaller studies published previously, they found a high incidence of maternal vascular malperfusion.

Unlike previous studies, this study evaluated the placenta following resolved SARS-CoV-2 infection. Interestingly, they found that placentas from pregnancies with resolved and active infection had maternal vascular malperfusion. They also tested for a specific pathological feature dependent on the gestational timing of infection, but did not find one. However, they did report a higher frequency of placental pathology findings if the infection occurred before 20 weeks of gestation.^22^

This study aimed to identify factors contributing to the development of placenta pathology following SARS-CoV-2 infection. We hypothesized that COVID-19 causes pathologic changes in the placenta that are not related to timing or severity of clinical disease. To test this hypothesis, we assembled a large cohort of patients diagnosed with COVID-19 during pregnancy whose placentas underwent comprehensive pathologic examination performed by an experienced placental pathologist blinded to demographics and outcomes data for each patient. Our study population includes patients of diverse racial and ethnic backgrounds, various gestational timing of infection, and a range of COVID-19 severity. The primary outcome we studied was a composite of placental pathology findings (high grade maternal vascular malperfusion (MVM) and/or presence of greater than 25 percent perivillous fibrin deposition), and exposures we compared included timing of infection during pregnancy (first or second trimester versus third trimester), severity of COVID-19, and SARS-CoV-2 variants (four time periods of the COVID-19 pandemic). Secondary outcomes included adverse pregnancy outcomes stratified by presence or absence of placental pathology.

## Methods

This study was carried out with Institutional Review Board approval (protocol #843277) using clinical and pathologic material collected from the Hospital of the University of Pennsylvania (HUP), Philadelphia, Pennsylvania.

### Selection of subjects

Women with a history of SARS-CoV-2 infection during pregnancy were identified either prospectively by a clinician/research coordinator present on the Labor and Delivery unit or retrospectively by searching the HUP pathology database for placentas with the clinical history of “COVID-19”, “COVID-19 positive”, “SARS-CoV-2”, “SARS-CoV-2 positive”, and “resolved COVID-19” from March 1, 2020 to February 28, 2022. During the study period, universal screening via nasopharyngeal polymerase chain reaction (PCR) for SARS-CoV-2 was performed for all admissions to the labor and delivery unit at our institution. The placentas of all patients with a positive SARS-CoV-2 test or a self-reported history of infection during pregnancy were sent for routine gross and microscopic pathologic examination. COVID-19 cases for this study were selected from this population based on the following criteria: singleton delivery at or after 28 weeks gestational age (third trimester), and a documented positive SARS-CoV-2 PCR test during pregnancy. Additionally, cases with fetal congenital or genetic anomalies or a known non-SARS-CoV-2 viral infection during this pregnancy (e.g., HIV, CMV, etc.) were excluded. In order to compare rates of placenta pathology between COVID-19 cases and healthy controls, we submitted placentas from a small group of healthy controls (N=26) who delivered during the same time period for routine gross and microscopic pathologic examination.

### Clinical and demographic data collection

Maternal clinical characteristics were extracted from electronic medical records including maternal age at delivery, gravidity, parity, race/ethnicity, pre-gravid body mass index (kg/m^2^), history of maternal disease (e.g. chronic hypertension, gestational hypertension, preeclampsia, pre-gestational diabetes, or gestational diabetes), and maternal use of aspirin during pregnancy. Obstetrical and neonatal outcomes that we studied included gestational age at delivery (weeks), preterm birth (before/after 37 weeks), birthweight, fetal sex, gestational hypertension, severe maternal morbidity,^23^ and severe neonatal morbidity.^24,25^ Birthweight z-scores were calculated using research calculator references and standardized by both gestational age and newborn sex.^26,27^ In accordance with the American College of Obstetricians and Gynecologists definition of severe maternal morbidity,^23^ the composite severe maternal morbidity outcome was defined as any (or multiple) of the following: (1) acute respiratory distress syndrome; (2) acute myocardial infarction; (3) amniotic fluid embolism; (4) congestive heart failure; (5) sepsis/shock; (6) hemorrhage; (7) eclampsia; (8) disseminated intravascular coagulation; (9) hysterectomy; (10) ICU admission; (11) acute renal failure; (12) assisted ventilation (intubation); and (13) death. There were no maternal deaths. In accordance with the Maternal-Fetal Medicine Units Network criteria for severe neonatal morbidity,^24,25^ the composite severe neonatal morbidity outcome was defined as any (or multiple) of the following: (1) respiratory distress syndrome; (2) transient tachypnea of the newborn; (3) sepsis; (4) assisted ventilation; (5) seizure; (6) grade 3 or 4 intraventricular hemorrhage; (7) necrotizing enterocolitis; (8) NICU admission; and (9) CPAP or supplemental oxygen.

The severity of maternal COVID19 disease was categorized based on National Institutes of Health and Society for Maternal-Fetal Medicine criteria initially issued in 2020:

1. *- Asymptomatic Infection*: Individuals who test positive for SARS-CoV-2 by virologic testing using a molecular diagnostic test but have no symptoms.
2. *- Mild Illness*: Individuals who have any of the various signs and symptoms of COVID 19 (e.g., fever, cough, sore throat, malaise, headache, muscle pain) without shortness of breath, dyspnea, or abnormal chest imaging.
3. *- Moderate Illness*: Individuals who have evidence of lower respiratory disease by clinical assessment or imaging and a saturation of oxygen (SpO_2_) ≥94% on room air at sea level.
4. *- Severe Illness*: Individuals who have respiratory frequency >30 breaths per minute, SpO_2_ <94% on room air at sea level, ratio of arterial partial pressure of oxygen to fraction of inspired oxygen (PaO_2_/FiO_2_) <300 mmHg, or lung infiltrates >50%
5. *- Critical Illness*: Individuals who have respiratory failure, septic shock, and/or multiple organ dysfunction.

The gestational age at the time of positive SARS-CoV-2 test was collected and the days from diagnosis to delivery were calculated. Maternal COVID-19 status at delivery was further categorized as active (positive test result at the time or within 14 days of delivery) or resolved (positive test result > 14 days before date of delivery). Infants delivered to patients with active COVID-19 at the time of delivery were routinely tested via nasopharyngeal swab PCR test for SARS-CoV-2 at 24 and 48 hours of life. If an infant was tested for SARS-CoV-2 after delivery, these results were also collected.

A publicly available surveillance sequencing dashboard of SARS-CoV-2 variants circulating in the Philadelphia region since March 2020 was utilized to categorize patients into one of four variant time periods (City of Philadelphia Department of Public Health, (https://microb120.med.upenn.edu/data/SARS-CoV-2/). As our institution is one of the main contributors to this dashboard and a majority of patient’s delivering at our institution are located within the geographic region covered by this collaborative effort, these data should represent our patient population. Based on the predominant variants present, patients were categorized based on the date of their positive SARS-CoV-2 PCR test as follows:

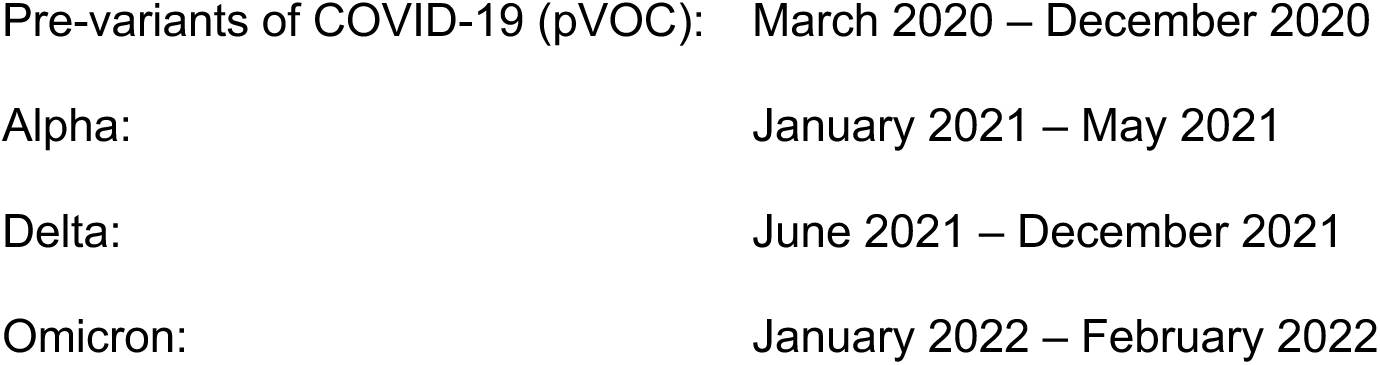

### Placenta pathologic data collection

All placentas were examined in the pathology department at HUP utilizing a systematic protocol, including recording the trimmed placental weight, membrane insertion site, gross appearance, dimensions of the placental disc, and the insertion, length, and diameter of the umbilical cord.

Histologic samples included sections of membranes, umbilical cord, and at least three full-thickness sections of non-lesion placental parenchyma. The placentas from COVID-19 cases were fixed in 10% buffered formalin for at least 24 hours prior to examination. Macroscopic and microscopic lesions were described primarily according to the Amsterdam Placental Workshop Group 2014 classification and further defined as per **Table 1**.^28–30^ A composite of high grade MVM and/or presence of greater than 25 percent perivillous fibrin deposition was the primary placental pathology outcome we investigated. Microscopic examination was performed by a single perinatal pathologist for all placentas (R.L.L.) blinded to exposures (i.e., timing and severity of COVID-19) and clinical outcomes. Subsequently, a second review on a subset of 32 placentas was performed to assess inter-observer reliability. Slides were reviewed independently by two pathologists (R.L.L. and J.C.G.) to assess for the presence or absence of: chronic inflammation including chronic villitis and chronic histiocytic intervillositis; fetal vascular malperfusion (FVM) including fetal vascular thrombi, avascular villi and villous stromal vascular karyorrhexis; MVM including decidual vasculopathy, villous infarction, accelerated villous maturation, distal villous hypoplasia; and increased perivillous fibrin deposition.

**Table 1.** Macroscopic and microscopic placental lesions (Amsterdam Placental Workshop Group 2014 classification).^24^ A composite of high grade maternal vascular malperfusion (MVM) or presence of greater than 25 percent perivillous fibrin deposition was the primary placental pathology outcome we analyzed.

| Variable | Lesions included | Definition of grading |
| --- | --- | --- |
| Chronic inflammation | <u>Membranes/chorionic plate:</u> chronic chorionitis, chronic amnionitis<br><u>Basal plate:</u> chronic deciduitis with plasma cells<br><u>Villi:</u> chronic villitis, intervillous plasma cells<br><u>Intervillous space:</u> chronic histiocytic intervillitis (≥2% involvement of intervillous space)<br><u>Fetal:</u> eosinophilic T-cell vasculitis | <u>High-grade:</u> ≥2 compartments with chronic inflammation<br><u>Low-grade:</u> 1 compartment with chronic inflammation<br><u>None:</u> no compartments with chronic inflammation<sup>23</sup> |
| Fetal vascular malperfusion (FVM) | Fetal vascular thrombi or intramural fibrin deposition in umbilical, chorionic, or stem villous vessel; at least 3 foci of avascular villi and/or villous stromal vascular karyorrhexis (≥3 villi/focus) | <u>High grade:</u> ≥2 fetal chorionic or stem villous vascular thrombi or a total of ≥45 avascular villi over 3 sections examined<br><u>Low grade:</u> any FVM lesions insufficient for a high-grade categorization<br><u>None:</u> no fetal vascular lesions<sup>24</sup> |
| Maternal vascular malperfusion (MVM) | <u>Decidual vasculopathy:</u> fibrinoid necrosis/acute atherosclerosis, persistent muscularization of basal plate arterioles, mural hypertrophy of membrane arterioles, basal decidual vascular thrombus, chronic decidual perivasculitis<br><u>Villous infarction</u><br><u>Accelerated villous maturation including increased syncytial knots for gestational age</u><br><u>Distal villous hypoplasia</u><br><u>Changes consistent with retroplacental hemorrhage/hematoma</u> | <u>High grade:</u> accelerated villous maturation and ≥1 of the following: placental weight <10<sup>th</sup> percentile, multiple villous infarcts, or distal villous hypoplasia<br><u>Low grade:</u> any MVM lesions insufficient for a high-grade categorization<br><u>None:</u> no MVM lesions<sup>25</sup> |
| Perivillous fibrin deposition | Increased perivillous fibrin in the intervillous space (more than expected for gestational age) | ≥10% perivillous fibrin deposition<br>≥25% perivillous fibrin deposition |

### Statistical Analysis

Statistical analyses were performed with SAS 9.4 for Windows. An alpha level of 0.05 was used to determine statistical significance; Bonferroni correction was used for multiple comparisons.

Timing of COVID-19 during pregnancy (first or second trimester versus third trimester) was the main exposure and a composite of placental pathology, defined as the presence of high grade MVM and/or presence of greater than 25 percent perivillous fibrin deposition, was the primary outcome.

Patient demographics included maternal age (years); race (Black/African American versus Caucasian/Asian/Other); gestational age (weeks); fetal sex (male versus female); chronic hypertension; diabetes; active infection at delivery; COVID-19 treatment; COVID-19 vaccination preceding COVID-19 diagnosis; COVID-19 severity (asymptomatic/mild versus moderate/severe/critical); COVID-19 variant (pVOC/alpha/delta/omicron); and infection exposure window (days from delivery).

Summary statistics were computed as frequencies and proportions for all categorical variables. Maternal age and exposure window were summarized with mean and standard deviation.

Chi-square test for independence, Student’s t-test, and Wilcoxon ranked sum test were performed to examine bivariable associations between timing of exposure and patient characteristics. All characteristics with P<0.05 were considered potential confounders and included in an adjusted logistic regression model.

Simple logistic regression was first performed to find the crude association between timing of COVID-19 during pregnancy (first or second trimester versus third trimester) and the composite placental pathology outcome. Next, potential confounding variables were placed in a regression model for adjusted estimates of the association between timing of infection and odds of the composite placental pathology outcome.

## Results

A total of 497 placentas from patients who had COVID-19 during pregnancy were evaluated for this study. Within this cohort, 156 patients had COVID-19 during the first or second trimester and 341 patients had COVID-19 during the third trimester (primary exposure). Sixty-three patients had the composite placental pathology outcome (high grade MVM and/or presence of greater than 25 percent perivillous fibrin deposition), and 434 did not have the primary outcome.

Demographic characteristics and secondary COVID-19 related exposures were compared between patients with COVID-19 in the first or second trimester versus patients with COVID-19 in the third trimester (timing of COVID-19 during pregnancy, primary exposure of interest, **Table 2**). Among the demographic characteristics we studied, maternal age was greater and patients were more likely to be multiparous among the group with COVID-19 in the first or second trimester. In accordance with study design, COVID-19 at delivery was less common and the interval from COVID-19 diagnosis to delivery was greater among the group with COVID-19 in the first or second trimester.

**Table 2.**
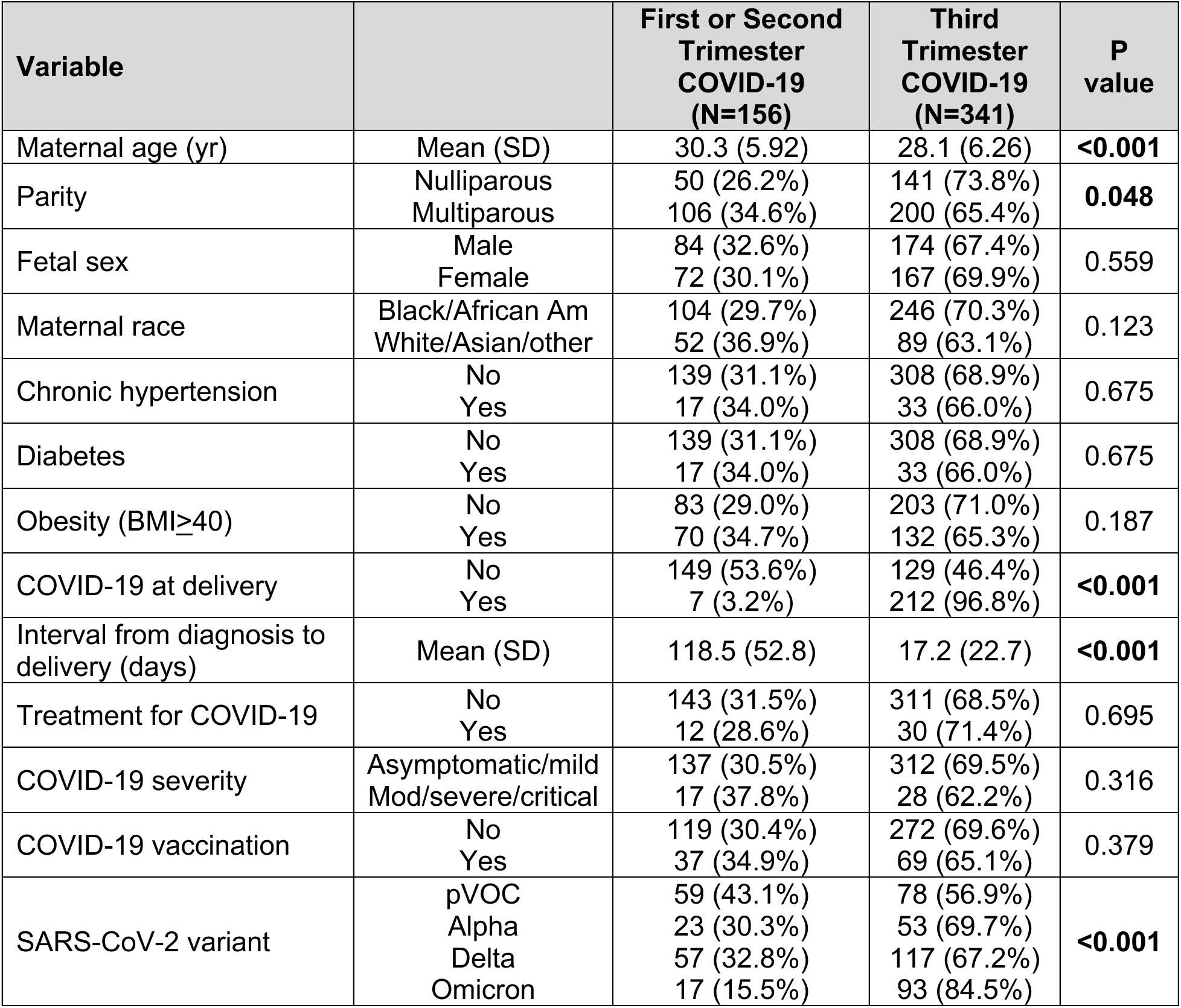
Demographics and secondary COVID-19 exposures comparing patients with COVID-19 in the first or second trimester and patients with COVID-19 in the third trimester (primary exposure). Chi-square and Fisher’s exact tests were used to find P-values for categorical variables. Student’s t-test and Wilcoxon ranked sum tests were used to find P-values for continuous measures. pVOC = pre-variants of COVID-19.

| Variable |  | First or Second Trimester COVID-19 (N=156) | Third Trimester COVID-19 (N=341) | P value |
| --- | --- | --- | --- | --- |
| Maternal age (yr) | Mean (SD) | 30.3 (5.92) | 28.1 (6.26) | **<0.001** |
| Parity | Nulliparous<br>Multiparous | 50 (26.2%)<br>106 (34.6%) | 141 (73.8%)<br>200 (65.4%) | **0.048** |
| Fetal sex | Male<br>Female | 84 (32.6%)<br>72 (30.1%) | 174 (67.4%)<br>167 (69.9%) | 0.559 |
| Maternal race | Black/African Am<br>White/Asian/other | 104 (29.7%)<br>52 (36.9%) | 246 (70.3%)<br>89 (63.1%) | 0.123 |
| Chronic hypertension | No<br>Yes | 139 (31.1%)<br>17 (34.0%) | 308 (68.9%)<br>33 (66.0%) | 0.675 |
| Diabetes | No<br>Yes | 139 (31.1%)<br>17 (34.0%) | 308 (68.9%)<br>33 (66.0%) | 0.675 |
| Obesity (BMI≥40) | No<br>Yes | 83 (29.0%)<br>70 (34.7%) | 203 (71.0%)<br>132 (65.3%) | 0.187 |
| COVID-19 at delivery | No<br>Yes | 149 (53.6%)<br>7 (3.2%) | 129 (46.4%)<br>212 (96.8%) | **<0.001** |
| Interval from diagnosis to delivery (days) | Mean (SD) | 118.5 (52.8) | 17.2 (22.7) | **<0.001** |
| Treatment for COVID-19 | No<br>Yes | 143 (31.5%)<br>12 (28.6%) | 311 (68.5%)<br>30 (71.4%) | 0.695 |
| COVID-19 severity | Asymptomatic/mild<br>Mod/severe/critical | 137 (30.5%)<br>17 (37.8%) | 312 (69.5%)<br>28 (62.2%) | 0.316 |
| COVID-19 vaccination | No<br>Yes | 119 (30.4%)<br>37 (34.9%) | 272 (69.6%)<br>69 (65.1%) | 0.379 |
| SARS-CoV-2 variant | pVOC<br>Alpha<br>Delta<br>Omicron | 59 (43.1%)<br>23 (30.3%)<br>57 (32.8%)<br>17 (15.5%) | 78 (56.9%)<br>53 (69.7%)<br>117 (67.2%)<br>93 (84.5%) | **<0.001** |

In unadjusted analyses, timing of COVID-19 during pregnancy was not associated with increased risk of the composite placental pathology outcome – 22/156 (14.1 percent) patients with COVID-19 in the first or second trimester compared to 41/341 (12.0 percent) with COVID- 19 in the third trimester (P=0.518, OR 1.20, 95 percent CI 0.69, 2.10; **Table 3**). Maternal age, parity, active infection at delivery, interval from time of diagnosis to delivery, and COVID-19 variant were associated with the primary exposure, trimester of infection (**Table 2**). Hence, in adjusted analyses, we controlled for these variables and determined timing of COVID-19 during pregnancy was not associated with odds of composite placental pathology in this sample (P=0.547, OR 1.33, 95 percent CI 0.53, 3.34).

**Table 3.** Unadjusted analyses of rates of composite placental pathology (primary outcome) compared between primary and secondary exposure groups. Chi-square tests were used to find P values. pVOC = pre-variants of COVID-19.

| Variable |  | Absence of composite placental pathology outcome (N=434) | Presence of composite placental pathology outcome (N=63) | P value |
| --- | --- | --- | --- | --- |
| Timing of COVID-19 | 1<sup>st</sup>/2<sup>nd</sup> trimester<br>3<sup>rd</sup> trimester | 134 (85.9%)<br>300 (88.0%) | 22 (14.1%)<br>41 (12.0%) | 0.518 |
| COVID-19 at delivery | No<br>Yes | 241 (86.7%)<br>193 (88.1%) | 37 (13.3%)<br>26 (11.9%) | 0.633 |
| Treatment for COVID-19 | No<br>Yes | 401 (88.3%)<br>32 (76.2%) | 53 (11.7%)<br>10 (23.8%) | **0.024** |
| COVID-19 severity | Asymptomatic/mild<br>Mod/severe/critical | 397 (88.4%)<br>35 (77.8%) | 52 (11.6%)<br>10 (22.2%) | **0.040** |
| SARS-CoV-2 variant | pVOC<br>Alpha<br>Delta<br>Omicron | 112 (81.8%)<br>68 (89.5%)<br>152 (87.4%)<br>102 (92.7%) | 25 (18.2%)<br>8 (10.5%)<br>22 (12.6%)<br>8 (7.3%) | 0.070 |
| COVID-19 vaccination | No<br>Yes | 337 (86.2%)<br>97 (91.5%) | 54 (13.8%)<br>9 (8.5%) | 0.144 |

Among the secondary COVID-19 related exposures we investigated, severity of disease (moderate/severe/critical versus asymptomatic/mild) and treatment for COVID-19 (yes/no) were associated with risk of the composite placental pathology outcome (P<0.05), while COVID-19 vaccination (yes/no) and COVID-19 active at delivery (yes/no positive SARS-CoV-2 PCR at delivery admission) were not associated with the composite placental pathology outcome.

Although there were not statistically different rates of the composite placental pathology outcome among the four SARS-CoV-2 variants (P=0.070, **Table 3**), we did observe a trend toward greater rates of the composite placental pathology outcome in the earlier epochs during the pandemic – pVOC (March 2020 – December 2020, 18.2 percent), Alpha (January 2021 – May 2021, 10.5 percent), Delta (June 2021 – December 2021, 12.6 percent), Omicron (January 2022 – February 2022, 7.3 percent).

After correcting for multiple comparisons (Bonferroni significance = 0.008 for analysis of six secondary outcomes), no secondary obstetrical/neonatal outcomes were associated with timing of COVID-19 during pregnancy (**Table 4**). In stratified analyses, we found that timing of COVID- 19 during pregnancy in patients with the composite placental pathology was not associated with risk of secondary maternal/neonatal outcomes, while timing of COVID-19 during pregnancy in patients without the composite placental pathology was associated with risk of spontaneous preterm birth (15/134 [11.2 percent] patients with COVID-19 in the first or second trimester versus 12/300 [4.0 percent] patients with COVID-19 in the third trimester; P=0.004, **Table 5**).

**Table 4.** Obstetrical and neonatal outcomes. SGA=small for gestational age (birthweight <10^th^ percentile, birthweight z-scores were calculated using research calculator references and standardized by both gestational age and newborn sex;^21,22^ SPTB=spontaneous preterm birth; PTB=preterm birth. P values were calculated by chi-square test; Bonferroni significance = 0.008 for analysis of six secondary outcomes.

| Outcome |  | First or Second Trimester COVID-19 (N=156) | Third Trimester COVID-19 (N=341) | P value |
| --- | --- | --- | --- | --- |
| SGA | BW>10<sup>th</sup> percentile | 139 (90.8%) | 287 (84.7%) | 0.062 |
|  | BW<10<sup>th</sup> percentile | 14 (8.2%) | 52 (15.3%) |  |
| Composite neonatal outcome | No | 123 (78.8%) | 274 (80.4%) | 0.698 |
|  | Yes | 33 (21.2%) | 67 (19.6%) |  |
| SPTB <37 wk | No | 140 (89.7%) | 326 (95.6%) | 0.012 |
|  | Yes | 16 (10.3%) | 15 (4.4%) |  |
| Medically indicated PTB <37 wk | No | 137 (87.8%) | 313 (91.8%) | 0.161 |
|  | Yes | 19 (12.2%) | 28 (8.2%) |  |
| Gestational hypertension | No | 121 (77.6%) | 253 (74.2%) | 0.419 |
|  | Yes | 35 (22.4%) | 88 (25.8%) |  |
| Severe maternal morbidity | No | 146 (93.6%) | 310 (90.9%) | 0.313 |
|  | Yes | 10 (6.4%) | 31 (9.1%) |  |

**Table 5.** Stratified analyses comparing risk of adverse pregnancy outcomes based on timing of COVID-19 during pregnancy (first or second trimester versus third trimester) in patients with or without the primary placental pathology outcome (high grade maternal vascular malperfusion (MVM) or presence of greater than 25 percent perivillous fibrin deposition). SGA=small for gestational age (birthweight <10^th^ percentile, birthweight z-scores were calculated using research calculator references and standardized by both gestational age and newborn sex;^21,22^ SPTB=spontaneous preterm birth; PTB=preterm birth. P values were calculated by chi-square test or Fisher’s exact test (if ≤5 samples in a cell).

| Patients with primary placental pathology outcome (N=63) |  |  |  |  |
| --- | --- | --- | --- | --- |
| Outcome |  | First or Second Trimester COVID-19 (N=22) | Third Trimester COVID-19 (N=41) | P value |
| SGA | BW>10<sup>th</sup> percentile | 17 (77.3%) | 25 (61.0%) | 0.265 |
|  | BW<10<sup>th</sup> percentile | 5 (22.7%) | 16 (39.0%) |  |
| Composite neonatal outcome | No | 13 (59.1%) | 28 (68.3%) | 0.465 |
|  | Yes | 9 (40.9%) | 13 (31.7%) |  |
| SPTB <37 wk | No | 21 (95.4%) | 38 (92.7%) | 1.000 |
|  | Yes | 1 (4.6%) | 3 (7.3%) |  |
| Medically indicated PTB <37 wk | No | 14 (63.6%) | 33 (80.5%) | 0.143 |
|  | Yes | 8 (36.4%) | 8 (19.5%) |  |
| Gestational hypertension | No | 14 (63.6%) | 26 (63.4%) | 0.986 |
|  | Yes | 8 (36.4%) | 15 (36.6%) |  |
| Severe maternal morbidity | No | 20 (90.9%) | 35 (85.4%) | 0.702 |
|  | Yes | 2 (9.1%) | 6 (14.6%) |  |
| Patients without primary placental pathology outcome (N=434) |  |  |  |  |
| Outcome |  | First or Second Trimester COVID-19 (N=134) | Third Trimester COVID-19 (N=300) | P value |
| SGA | BW>10th percentile | 122 (93.1%) | 262 (87.9%) | 0.105 |
|  | BW<10th percentile | 9 (6.9%) | 36 (12.1%) |  |
| Composite neonatal outcome | No | 110 (82.1%) | 246 (82.0%) | 0.982 |
|  | Yes | 24 (17.9%) | 54 (18.%) |  |
| SPTB <37 wk | No | 119 (88.8%) | 288 (96.0%) | 0.004 |
|  | Yes | 15 (11.2%) | 12 (4.0%) |  |
| Medically indicated PTB <37 wk | No | 123 (91.8%) | 280 (93.3%) | 0.564 |
|  | Yes | 11 (8.2%) | 20 (6.7%) |  |
| Gestational hypertension | No | 107 (79.8%) | 227 (75.7%) | 0.339 |
|  | Yes | 27 (20.2%) | 73 (24.3%) |  |
| Severe maternal morbidity | No | 126 (94.0%) | 275 (91.7%) | 0.391 |
|  | Yes | 8 (6.0%) | 25 (8.3%) |  |

Timing of COVID-19 during pregnancy and secondary COVID-19 exposures were not associated with other individual placental pathology outcomes that we investigated (**Table 6**, P>0.05 for all outcomes). In order to compare rates of placenta pathology between COVID-19 cases and healthy controls, we analyzed placentas from a small group of healthy controls (N=26) who delivered during the same time period (March 1, 2020 to February 28, 2022).

**Table 6.** Secondary placenta pathology outcomes.

| Outcome | Exposure | Unadjusted P value | Bonferroni P value<sup>1</sup> |
| --- | --- | --- | --- |
| High grade fetal vascular malperfusion | Timing of COVID-19 (trimester) | 0.580 | 1 |
|  | COVID-19 at delivery | 0.812 | 1 |
|  | Treatment for COVID-19 | 0.7518 | 1 |
|  | COVID-19 severity | 0.180 | 0.901 |
|  | SARS-CoV-2 variant | 0.186 | 0.932 |
| Fetal vascular thrombi | Timing of COVID-19 (trimester) | 0.554 | 1 |
|  | COVID-19 at delivery | 0.913 | 1 |
|  | Treatment for COVID-19 | 0.453 | 1 |
|  | COVID-19 severity | 0.784 | 1 |
|  | SARS-CoV-2 variant | 0.342 | 1 |
| High grade chronic inflammation | Timing of COVID-19 (trimester) | 0.303 | 1 |
|  | COVID-19 at delivery | 0.419 | 1 |
|  | Treatment for COVID-19 | 0.917 | 1 |
|  | COVID-19 severity | 0.100 | 0.501 |
|  | SARS-CoV-2 variant | 0.898 | 1 |
| Chronic histiocytic intervillitis or high grade chronic villitis | Timing of COVID-19 (trimester) | 0.592 | 1 |
|  | COVID-19 at delivery | 0.196 | 0.982 |
|  | Treatment for COVID-19 | 0.965 | 1 |
|  | COVID-19 severity | **0.014** | 0.069 |
|  | SARS-CoV-2 variant | 0.926 | 1 |

<sup>1</sup>Bonferroni adjustment for five potential predictors to minimize chances of a Type 1 error.

Although low grade MVM or FVM was observed in 7/26 controls (26.9 percent), none of the placentas from 26 healthy controls had the composite placental pathology outcome (high grade MVM or presence of greater than 25 percent perivillous fibrin deposition).

The previously collected data were not changed by the results of our interobserver reliability assessment. Interobserver agreement between the two pathologists ranged from substantial to almost perfect for the two components of the composite placental pathology outcome: kappa = 0.761 for >25 percent perivillous fibrin deposition, kappa = 0.930 for high grade MVM.^31^

## Discussion

In this large cohort, the composite placental pathology outcome (high grade MVM or presence of greater than 25 percent perivillous fibrin deposition) was relatively common among COVID-19 cases but was unrelated to timing of COVID-19 during pregnancy (first or second trimester versus third trimester) and adverse pregnancy outcomes. Among the secondary COVID-19 related exposures we investigated, severity of disease (moderate/severe/critical versus asymptomatic/mild) and treatment for COVID-19 (yes/no) were associated with risk of the composite placental pathology outcome. Spontaneous preterm birth occurred more frequently among patients with COVID-19 in the first or second trimester compared to COVID-19 in the third trimester, but in stratified analyses, we discovered that increased rates of spontaneous preterm births were restricted to patients with COVID-19 in the first or second trimester who did not have the composite placental pathology. An interesting trend we observed is that the composite placental pathology outcome occurred most frequently among pregnant patients who had COVID-19 early in the pandemic: March – December 2020 (pre-variants of COVID-19, 25/137, 18.2 percent) versus January – February 2022 (Omicron, 8/110, 7.3 percent).

Early reports during the COVID-19 pandemic showed low rates of placental infection with SARS-CoV-2 but high rates of placental pathology, including MVM, FVM, and villitis of unknown origin.^13–17^ A later meta-analysis that included 56 studies and pathological examination of 1,008 placentas from patients who had COVID-19 largely at the time of delivery revealed rates of MVM, FVM, and perivillous fibrin deposition that ranged from 25 to 33 percent, and these rates were higher in cases compared to healthy controls who did not have COVID-19 during pregnancy.^32^ Placental pathology is extensive in pregnancies complicated by acute SARS-CoV-2 infection, although less is known about placental pathology following a resolved infection during pregnancy. Here we report similar placental pathology in pregnancies with a resolved SARS-CoV-2 infection, even months after infection. This confirms the previous report by Corbetta-Rastelli et al in a smaller cohort.^22^

While asymptomatic infections cause placental pathology, it has previously been reported that pathology is independent of severity of disease.^32^ This conclusion is limited by the small cohorts reported thus far. In our cohort, we discovered that patients with a moderate, severe, or critical clinical presentation have a higher incidence of the composite placental pathology outcome than patients with an asymptomatic or mild case. We propose the difference between our study and others is due to the large sample size and assessment by the same team of pathologists.

Placental pathology is not correlated with adverse pregnancy outcomes. Other investigators studying COVID-19 in Philadelphia found a lack of correlation between preterm birth and placental pathology and suggested placental pathology and preterm birth are caused by divergent pathways.^33^ We found that the risk of preterm birth is highest in COVID-19 patients without the composite placental pathology. These investigators also reported higher rates of placental vascular pathology in patients with COVID-19 but not a COVID-19 vaccine during pregnancy compared to controls.^33^ We found a trend toward higher rates of the composite placenta pathology in patients who did not receive the COVID-19 vaccine compared to patients who did receive the COVID-19 vaccine (P=0.144), but the unvaccinated cases probably occurred earlier in the pandemic when the composite placental pathology outcome was observed most frequently.

The most recent studies of placental pathology in COVID-19 cases demonstrate high rates of placental pathology, and these investigators recommend that COVID-19 is a risk factor that requires close monitoring during pregnancy.^34,35^ Our data do not support these recommendations. We found that the composite placental pathology was unrelated to adverse pregnancy outcomes, and patients with COVID-19 early in pregnancy did not have higher rates of small for gestational age neonates (birthweight <10^th^ percentile), preterm birth, the composite neonatal outcome, gestational hypertension, or severe maternal morbidity compared to patients with COVID-19 during the third trimester. Hence, we do not believe that uncomplicated COVID- 19 during pregnancy requires intensive fetal surveillance or detailed pathological examination of placentas after delivery.

The reasons for high-rates of placental pathology in COVID-19 cases (63/497 [12.7 percent] in our cohort had high grade MVM or presence of greater than 25 percent perivillous fibrin deposition) remain poorly understood. COVID-19 induces systemic pro-inflammatory and pro- coagulant effects, so future work may focus on inflammatory mediators and coagulation factors that affect the placenta in COVID-19 cases.

Our study had several notable strengths and limitations. We believe this is the largest single center cohort of COVID-19 cases during pregnancy in which placental pathology was examined in detail. The study population is representative of COVID-19 cases in large medical centers that employed universal SARS-CoV-2 screening for patients admitted to labor and delivery units during the pandemic, and careful clinical phenotyping was performed. The primary pathologist was blinded to COVID-19 factors (i.e., timing and severity of disease) and pregnancy outcomes. Unfortunately, the study was limited by the small number of unaffected control placentas that were evaluated during the pandemic, because placentas from healthy patients with normal pregnancy outcomes are not routinely sent to pathologists for evaluation. In addition, the primary exposure we evaluated (COVID-19 during the first or second trimester) occurred in only 156 of the total 497 patients within the overall cohort.

We conclude that placental vascular abnormalities occur in a large number of COVID-19 cases but do not appear to be related to timing of COVID-19 during pregnancy. However, COVID-19 severity and maternal race influence the incidence of placental pathology. In stratified analyses among patients with COVID-19 early in pregnancy, the composite placental pathology outcome was not associated with higher rates of severe maternal morbidity or the composite adverse neonatal outcome. Hence, we do not believe that intensive fetal surveillance (i.e., serial non-stress tests or biophysical profiles) and detailed examination of placental pathology are warranted in otherwise uncomplicated cases of COVID-19 during pregnancy. Additional research is needed to understand why these high rates of placental vascular abnormalities occur in COVID-19 cases.

## Data Availability

All data produced in the present study are contained in the manuscript or available upon reasonable request to the authors

